# Automated segmentation of brain metastases in T1-weighted contrast-enhanced MR images pre and post Stereotactic Radiosurgery

**DOI:** 10.1101/2023.07.07.23292387

**Authors:** Hemalatha Kanakarajan, Wouter De Baene, Patrick Hanssens, Margriet Sitskoorn

## Abstract

**Background and purpose:** Accurate segmentation of brain metastases on Magnetic Resonance Imaging (MRI) is a tedious and time-consuming task for radiologists that could be optimized with deep learning (DL) methods. Previous studies that evaluated the performance of several DL algorithms focused on training and testing the models on the planning MRI only. The purpose of this study is to evaluate well-known DL approaches (nnU-Net and MedNeXt) for their performance on both planning and follow-up MRI.

**Materials and methods:** Pre-treatment brain MRIs were collected retrospectively for 263 patients at the Gamma Knife Center of Elisabeth-TweeSteden Hospital (ETZ). The patients were split into 203 patients for training and 60 patients for testing. For these 60 patients, the follow-up MRIs were also retrospectively collected. To increase heterogeneity, we added the publicly available MRI from the Mathematical oncology laboratory for 75 patients to the training data. The performance was compared between the two models, with and without addition of the public data.

**Results:** All models obtained a good Dice Similarity Coefficient (all DSC >= 0.928) for planning MRI. MedNeXt trained with combined data was the best performing model for follow-ups at 3, 15 and 21 months (DSC of 0.796, 0.725, and 0.720 respectively) while nnU-Net trained with combined data was the best performing model for follow-ups at 6, 9 and 12 months (DSC of 0.738, 0.740 and 0.713 respectively).

**Conclusion:** The models achieved a good performance score for planning MRI. Though the models performed worse for follow-ups, addition of public data enhanced their performance, providing a viable solution to improve their efficacy for the follow-ups. There was only limited difference in performance of the two algorithms. These algorithms hold promise as a valuable tool for clinicians for automated segmentation of pre- and postsurgical MRI during treatment planning and response evaluations, respectively.

## Background

Brain metastases (BM) are the predominant intracranial tumors seen in adults [1]. It is estimated that about one-fifth of all cancer patients will ultimately develop BM [2]. Advancements in primary tumor treatments have increased life expectancy and hence the probability of developing BM [1]. The presence of BM is associated with a substantial increase in morbidity and mortality rates among cancer patients [3]. Stereotactic Radiosurgery (SRS) is a treatment option in which the BM are targeted very precisely, whereby the dose of radiation to the healthy brain tissue is limited. SRS has emerged as a well accepted treatment modality in the current standard of care for the treatment for BM [4].

For SRS treatment planning and treatment response evaluation, the physician must manually delineate numerous lesions on three-dimensional Magnetic Resonance Imaging (MRI) or Computed Tomography (CT) scans. This manual process is labor-intensive and prone to considerable variability among physicians [5]. Introducing an automatic and reliable system for detecting and delineating BM could facilitate more precise treatment delivery in the radiotherapy clinic. Automated tools that assist radiologists and radiation oncologists can positively influence both efficiency and efficacy in detecting and delineating multiple metastases.

Deep learning (DL) models have shown great promise in medical image analysis, particularly in detection, segmentation and classification tasks with the potential to improve clinical workflow [6]. There are plenty of studies on automated segmentation of primary tumors using DL algorithms [7,8,9,10,11]. Several approaches have also been introduced for BM segmentation on MRI using DL [12]. In 2015, Losch et al. [13] produced state-of-the-art results in automated segmentation of BM on MRI using deep convolutional networks. Since then, a large variety of network architectures for DL such as Convolutional Neural Networks (CNNs) [14] and DeepMedic [15] have been tested. However, a notable limitation of these studies is their exclusive focus on training and testing the models solely on the planning MRI (e.g. [14,16, 17, 18, 19, 20, 21]), potentially overlooking variations in the performance of the DL algorithms when applied to follow-up MRI. Such discrepancies may arise due to radiation-induced shrinkage of the tumors. It is imperative to assess the performance of DL algorithms on the follow-up MRI to ascertain their utility in assisting the clinicians in the response evaluation during follow-ups. Jalalifar et al. [22] evaluated the performance of a DL model on follow-up MRI but their study provided the performance results for only five sample patients.

One of the popular DL network architectures is the so-called nnU-Net [23]. Isensee et al. [24] demonstrated how this architecture achieved state of the art performance on different challenges in medical image segmentation by applying it to 10 international biomedical image segmentation challenges comprising 19 different datasets and 49 segmentation tasks across a variety of organs, organ substructures, tumors, lesions and cellular structures in MRI, CT and electron microscopy images. Ziyaee et al. [25] evaluated the effectiveness of this algorithm specifically for segmentation of BM by training and testing it with planning MRI only. The model achieved an overall Dice Similarity Coefficient (DSC) of 82.2%, which shows good segmentation performance. Recently, the transformer technique [26] has emerged as a noteworthy alternative to traditional CNNs in the medical domain, being employed for various tasks like classification, detection, and segmentation [27, 28, 29]. This has posed a significant challenge to existing CNN-based solutions [30] like nnU-Net.

Both approaches, however, have their own advantages and limitations. CNNs can accurately segment tumors by analyzing local details in the images [31]. But, CNN-based approaches generally exhibit limitations for modeling long-range dependencies. On the other hand, transformers are effective in considering the broader context of the entire images, but can result in limited localization abilities due to lack of detailed localization information [32]. Some network architectures now incorporate both convolutional layers and transformers to leverage the strengths of both approaches, aiming for improved performance and overcoming the limitations of each individual architecture [33]. An example is ConvNeXt [34] which combines the strenghts of both approaches. Building upon this, Roy et. al [35] introduced MedNeXt, a modernised and scalable convolutional architecture customised to challenges of data-scarce medical settings.

Compared to other algorithms, the nnU-Net and MedNeXt algorithms achieved better segmentation performance [35]. MedNeXt achieved state-of-the-art performance benefits on segmentation tasks of varying modality and sizes and hence Roy et al. [35] proposed MedNeXt as a strong and modernized alternative to standard ConvNets like nnU-Net for building deep networks for medical image segmentation. MedNeXt achieved this performance against baselines consisting of Transformer-based, convolutional and large kernel networks. However, the effectiveness of the nnU-Net and MedNeXT algorithms specifically for the segmentation of the BM follow-up images has not yet been evaluated.

The present study aims to bridge this gap by assessing the applicability of these state-of-the-art algorithms for automated segmentation of both planning and follow-up BM images. Typically, in most hospitals, segmentation of BM is performed solely on the planning MRI scans and not on the follow-up scans. This absence of ground truth (GT) segmentations poses a challenge for training DL algorithms with follow-up scans. In this research, we evaluated the performance of the nnU-Net and MedNeXt algorithms by training them with planning images and publicly available segmented images, then testing them on both planning and follow-up images. We used publicly available BM images and added these images to the training data to increase the heterogeneity of the training data. This evaluation will help to understand whether these state-of-the-art DL algorithms can assist the clinicians in detection and segmentation of BM images for treatment planning and treatment response evaluation during follow-ups.

## Method

For this study, pre-treatment contrast-enhanced (with triple dose gadolinium) T1-weighted brain MRIs of 263 BM patients were used. Scans were made as part of clinical care at the Gamma Knife Center of the Elisabeth-TweeSteden Hospital (ETZ) between 2015 and 2021 at Tilburg, The Netherlands. These planning MRI scans were collected using a 1.5T Philips Ingenia scanner (Philips Healthcare, Best, The Netherlands). The voxel size was 0.82 × 0.82 × 1.5mm3. The total of 263 patients were split into 203 patients for model training and 60 patients for testing. For the 203 patients in the training data set, the treatment type was guided by the tumor volume assessed on the planning MRI. The patients underwent either Gamma Knife Radiosurgery (GKRS) at the Gamma Knife Center or were referred to WBRT or surgery at the other departments. The 60 patients who were part of the testing set are from the set of patients included in the Cognition And Radiation Study A (CAR-Study A) at ETZ [36]. Our test set is a random subset of the set of the patients included in this CAR-Study A. All the patients in the test data set underwent GKRS. Patients with other brain tumor types (e.g. meningioma) in addition to BM were excluded from the test data set (n=6). After this exclusion, there were 54 patients in the test data set. For all patients in the training and test data set, the segmentations of the baseline GT were manually delineated by expert oncologists and neuroradiologists at ETZ. The manually delineated GT for follow-up scans were only available for the patients who were part of the CAR-Study A.

For the 54 patients used for testing, the post treatment contrast enhanced (with single dose gadolinium) T1-weighted follow-up MRI scans were also retrospectively collected. Though the slice thickness of the follow-up scans ranged from 0.21 mm to 1.5 mm, the majority of the scans had a slice thickness of 0.8 mm. The voxel size for all scans for the x- and y-dimension was 0.79 and 0.78 mm, respectively. The images from 6 follow-up (FU) sessions were available. The FU scans were made at 3, 6, 9, 12, 15, and 21 months after treatment. For these follow-ups, scans of 54(FU1), 41(FU2), 32(FU3), 27(FU4), 19(FU5) and 14(FU6) patients were available.

As a first preprocessing step, all the MRI scans were registered to standard MNI space using Dartel in SPM12 (Wellcome Trust Center for Neuroimaging, London, UK), implemented in Python (version 3.11) using the Nipype(Neuroimaging in Python–Pipelines and Interfaces) software package (version 1.8.6) [37]. The voxel size of the normalized image was set to 1*1*1 mm^3^. For all other normalization configurations, the default values offered by SPM12 were used. One other preprocessing step was to combine the GT labels for patients with more than one BM in one single GT mask. FSL library (Release 6.0) was used for this integration[38].

We also used the publicly available BM images from the Mathematical oncology laboratory provided by Ocaña-Tienda et al. [39] and added these images to the training data for models which were trained with the combination of ETZ data and public data. This data set contained 355 contrast-enhanced (with single dose of contrast) T1-weighted planning and follow-up MRI for 75 patients. The voxel size for all scans for the x- and y-dimensions ranged from 0.39 mm to 1.01 mm. The median slice thickness is 1.30 mm. Similar to the scans from ETZ, all the MRI scans from this public data set were also registered to standard MNI space with a voxel size of 1*1*1 mm^3^.

The nnU-Net algorithm, a framework built on top of the U-Net [23], makes key design decisions regarding pre-processing, post-processing, data augmentation, network architecture, training scheme, and inference, all tailored to the specific properties of the dataset at hand [23]. It analyzes the provided training cases and automatically configures a matching U-Net-based segmentation pipeline. These automatic design choices allow nnU-Net to perform well on many medical segmentation tasks. nnU-Net readily executes systematic rules to generate DL methods for previously unseen datasets without the need for further optimization [23]. The nnU-Net (verison 1) model was trained with the planning MRI in 3d full resolution mode. The trained model was then tested separately against the planning and follow-up images.

On the other hand, MedNeXt represents a novel approach to medical image segmentation, drawing inspiration from transformers. The architecture of MedNeXt includes ConvNeXt blocks, which are used for processing the image data. These blocks help in efficient sampling of the image [35]. MedNeXt also uses a novel technique to adjust the size of the processing units (kernels). MedNeXt is also customized to the challenges of sparsely annotated medical image segmentation datasets and is an effective modernization of standard convolution blocks for building deep networks for medical image segmentation[35]. MedNeXt offers 4 predefined architecture sizes (Small, Base, Medium, Large) and 2 predefined kernel sizes (3*3*3, 5*5*5). The combination that we used for training the model was Small with 3*3*3 kernel size.

The different models that we created are,

1. nnU-Net trained with ETZ BM data only (n=203).
2. nnU-Net trained with ETZ BM data and BM public data (n=558).
3. MedNeXt trained with ETZ BM data only (n=203).
4. MedNeXt trained with ETZ BM data and BM public data (n=558).

We evaluated the performance of these models on both planning and follow-up MRI. To assess the quality of the resulting segmentations, multiple metrics were employed. The DSC measures the overlap with the GT (ranging from 0 for no overlap to 1 for perfect overlap) per patient. It is calculated by dividing the double of the area of overlap by the sum of the areas of the predicted and the GT segmentation. The algorithm’s performance in detecting individual metastases was measured by sensitivity (number of voxels in the detected metastases divided by the number of voxels in all metastases contained in GT), and by the False Negative Rate (FNR). The FNR is the probability that a true metastasis will be missed by the model. In the results section, these metrics are presented for the predictions done for baseline and for the follow up test data.

## Results

Table 1 shows the characteristics of patients from the ETZ and public data set included in our study.

**Table 1:**
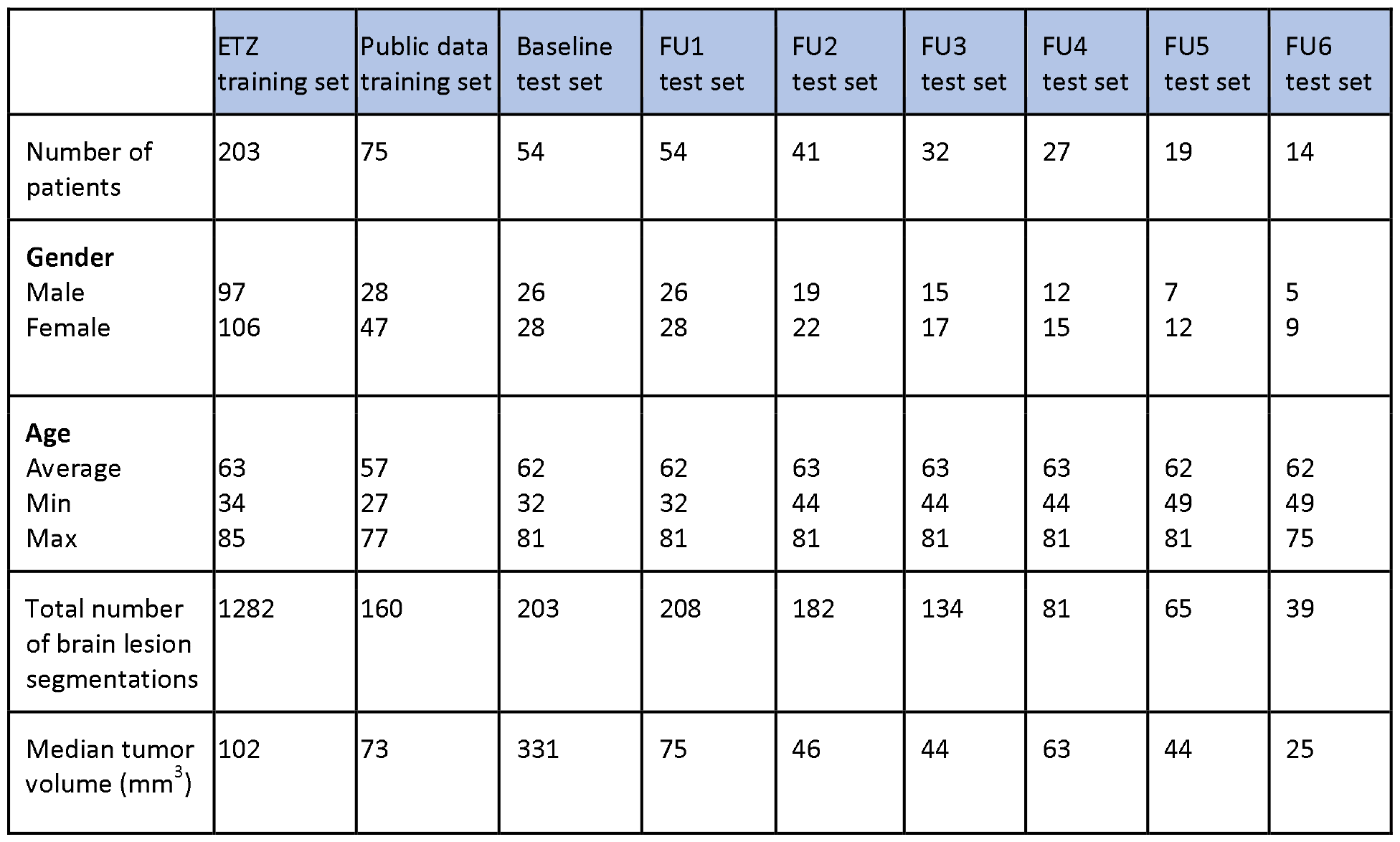
Characteristics of patients from ETZ and public data set.

The DSC obtained for the baseline and the FU tests for the four models are shown in table 2 and visually depicted in Figure 1.

**Table 2:** DSC of the segmentation models. In bold is the value of the best DSC for the corresponding test set.

| DSC | Baseline | FU1 | FU2 | FU3 | FU4 | FU5 | FU6 |
| --- | --- | --- | --- | --- | --- | --- | --- |
| Model 1 – nnU-Net trained with ETZ data only | **0.940** | 0.759 | 0.667 | 0.604 | 0.589 | 0.666 | 0.574 |
| Model 2 - nnU-Net trained with ETZ and public data | 0.928 | 0.784 | **0.738** | **0.740** | **0.713** | 0.724 | 0.704 |
| Model 3 - MedNeXt trained with ETZ data only | **0.940** | 0.762 | 0.692 | 0.680 | 0.606 | 0.672 | 0.707 |
| Model 4 - MedNeXt trained | **0.940** | **0.796** | 0.730 | 0.725 | 0.672 | **0.725** | **0.720** |
| with ETZ and public data |  |  |  |  |  |  |  |

**Figure 1:**
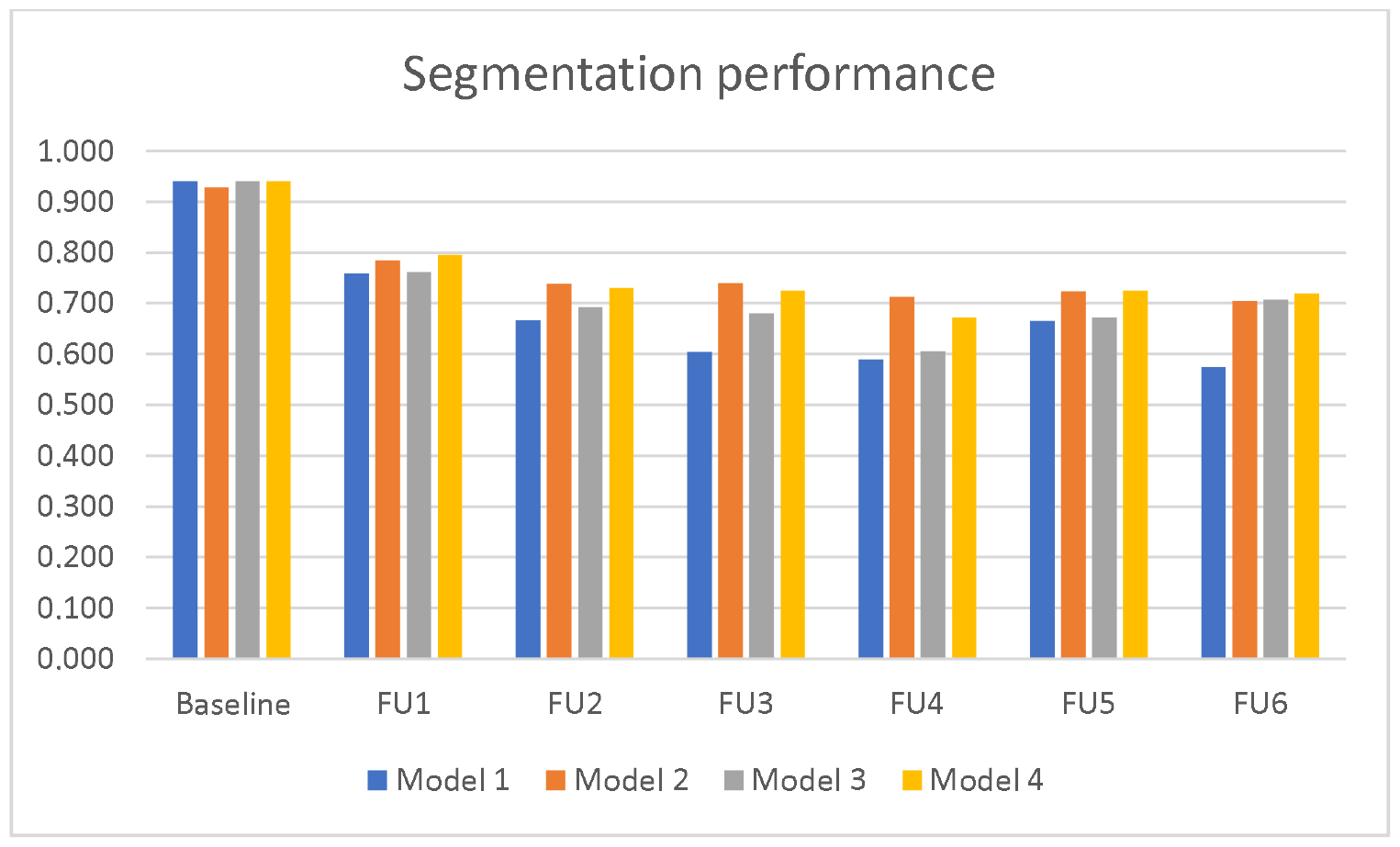
DSC of the segmentation models

All four models obtained a good DSC for planning MRI. The models 1, 3 and 4 had the same DSC of 0.940 for the planning MRI. For model 2, we obtained a slightly lower DSC of 0.928.

All four models obtained a lower DSC for the segmentation of follow-up MRI when compared with the DSC of planning MRI. Model 4 (MedNeXt trained with both ETZ and public data) had a DSC of 0.796 for FU1. This is higher than the DSC of other models for FU1. Similarly, model 4 had the highest DSC for FU5 and FU6. Model 2 (nnU-Net trained with both ETZ and public data) had a DSC of 0.738, 0.740 and 0.713 for FU2, FU3 and FU4 respectively. This is higher than the DSC of other models for FU2, FU3 and FU4.

The models which included the public data also in the training data set (model 2 and 4) performed better for the follow-ups when compared to the models which were trained with ETZ data only (model 1 and 3). For FU1, model 4 had a DSC of 0.796 for FU1. This is 0.034 higher than model 3 (MedNeXt trained with ETZ data only). Similarly, for FU5 and FU6, the model 4 had a DSC which is 0.053 and 0.013 respectively higher than the DSC of model 3. For FU2, FU3 and FU4, model 2 had a DSC of 0.071, 0.136 and 0.124 respectively higher than the DSC of model 1 (nnU-Net trained with ETZ data only).

The FNR and the sensitivity obtained for the baseline and the FU tests for the four models are shown in table 3 and table 4 respectively.

**Table 3:** FNR of the segmentation models. In bold is the value of the best FNR for the corresponding test set.

| FNR | Baseline | FU1 | FU2 | FU3 | FU4 | FU5 | FU6 |
| --- | --- | --- | --- | --- | --- | --- | --- |
| Model 1 – nnU-Net trained with ETZ data only | 0.066 | 0.289 | 0.379 | 0.446 | 0.471 | 0.410 | 0.488 |
| Model 2 - nnU-Net trained with ETZ and public data | 0.063 | 0.081 | 0.164 | 0.158 | **0.183** | **0.168** | 0.168 |
| Model 3 - MedNeXt trained with ETZ data only | 0.066 | 0.218 | 0.332 | 0.324 | 0.416 | 0.344 | 0.342 |
| Model 4 - MedNeXt trained with ETZ and public data | **0.063** | **0.074** | **0.157** | **0.155** | 0.220 | 0.183 | **0.162** |

**Table 4:**
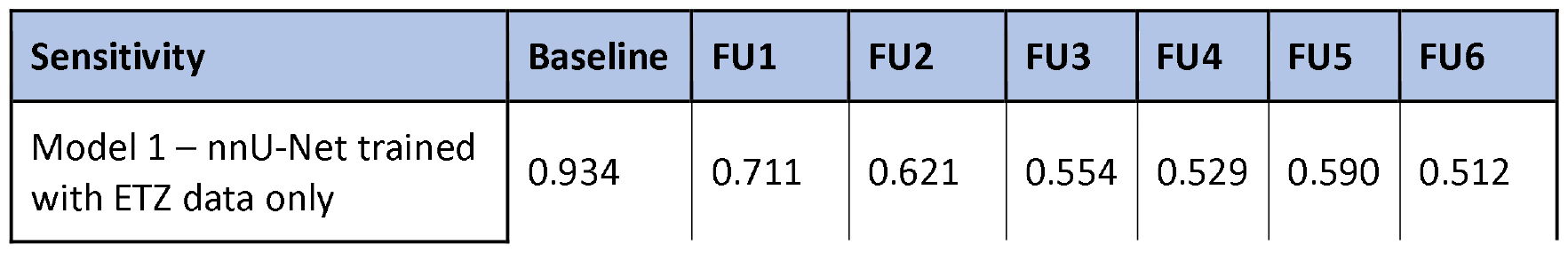
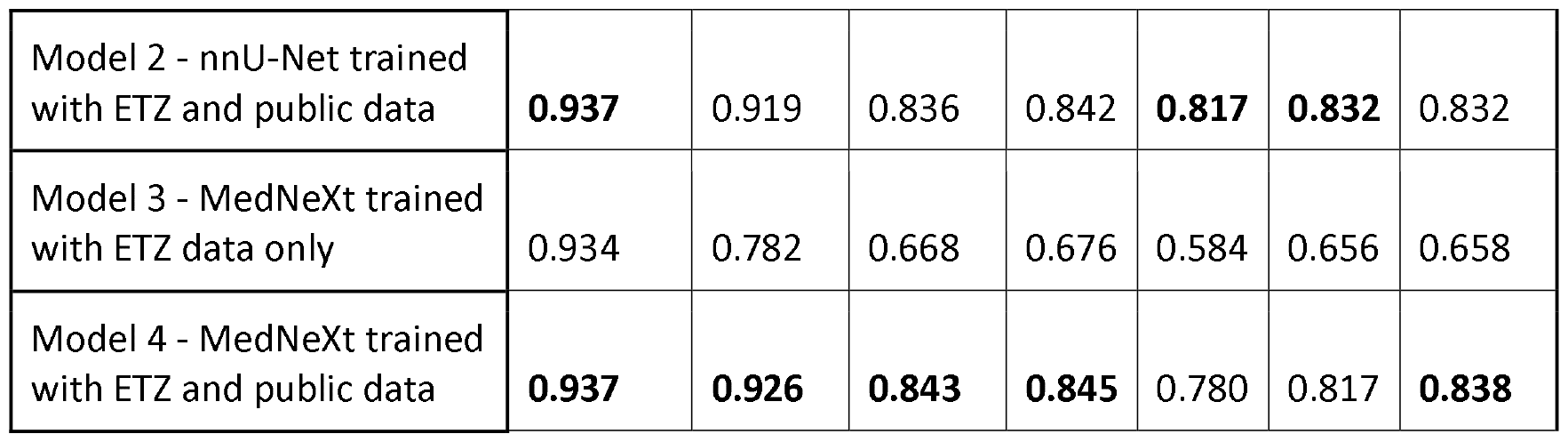
Sensitivity of the segmentation models. In bold is the value of the best sensitivity for the corresponding test set.

| Sensitivity | Baseline | FU1 | FU2 | FU3 | FU4 | FU5 | FU6 |
| --- | --- | --- | --- | --- | --- | --- | --- |
| Model 1 – nnU-Net trained with ETZ data only | 0.934 | 0.711 | 0.621 | 0.554 | 0.529 | 0.590 | 0.512 |
| Model 2 - nnU-Net trained with ETZ and public data | **0.937** | 0.919 | 0.836 | 0.842 | **0.817** | **0.832** | 0.832 |
| Model 3 - MedNeXt trained with ETZ data only | 0.934 | 0.782 | 0.668 | 0.676 | 0.584 | 0.656 | 0.658 |
| Model 4 - MedNeXt trained with ETZ and public data | **0.937** | **0.926** | **0.843** | **0.845** | 0.780 | 0.817 | **0.838** |

These results show that Model 2 and Model 4 had a superior FNR and sensivity when compared to the other two models.

Another interesting finding is that the models also detected some tumors that were missing in the GT. Some of the extra tumors detected by the models were part of the GT of subsequent scans. For example, for some patients the models detected an extra tumor in the baseline test which was not containted in the GT masks but which was part of the FU1 GT masks.

## Discussion

In the present work we assessed the effectiveness of the nnU-Net and MedNeXt algorithms for automated segmentation of both planning and follow-up MRI for BM patients. We conducted experiments by training four distinct models using these algorithms. Specifically, two nnU-Net models were trained – one solely with ETZ data and the other with the combination of ETZ and public data. Similarly, two MedNeXt models were trained – one exclusievely with ETZ data and the other with the combination of ETZ and public data.

As shown in Table 2, three of the models achieved a DSC of 0.940 on planning MRI while Model 2 (nnU-Net trained with ETZ and public data) achieved a DSC of 0.928. These results indicate a good performance of the models in segmenting BM on the planning MRI, with minimal variation in performance across the different models. Notably, the performance of our models for the planning MRI surpassed that reported in similar studies. For example, Hsu et al. [19] expounded a fully 3D DL approach capable of automatically detecting and segmenting BM on MRI and on CT scans. The DSC of this algorithm was found to be 0.76. Grøvik et al. [21] observed a DSC of 0.79 while evaluating a DL algorithm for detection and segmentation of BM on multisequence MRI. Ziyaee et al. [25] evaluated the effectiveness of the nnU-Net algorithm specifically for segmentation of BM by training and testing it with planning MRI only and achieved an overall DSC of 0.82.

However, during the evaluation on the planning MRI, our models exhibited some limitations. They tended to miss metastases situated near a blood vessel or the tentorium, while also producing some false positive segmentations by sometimes identifying blood vessels as BM. However the models did detect and segment some tumors that were missed in the GT. Some of these extra tumors that were detected by the models were part of the GT of the subsequent follow-up scans. This suggests the potential utility of these models in assisting clinicians with the early detection and segmentation of the tumors.

In contrast to the robust performance on planning MRI, the models showed lower efficacy for the follow-up images. This decline could be attributed to several factors, including the radiation effect which causes the tumors to shrink over time. Table 1 shows that the median tumor volume of the follow-up images is less than the planning MRI and also the median tumor volume decreased on subsequent follow-ups. The detection and segmentation performances of the DL algorithms tend to decrease for smaller lesions [20]. Hence, the shrinkage of the tumors over time due to the radiation effect could be a reason for the lower performance for successive follow-up scans. The decrease in performance for the follow-ups may also be due to the different dose of contrast administered during follow-up scans compared to planning scans. The planning MRI were contrast-enhanced with triple-dose gadolinium and the follow-up images were contrast-enhanced with single-dose gadolinium. Additionally, changes in tumor texture over successive follow-up scans, after multiple sessions of treatment, and variations in slice thickness between the planning and follow-up images might contribute to the diminished performance of the models on the follow-up MRI. The experiments of You et al. [40] confirmed that the performance of DL models change due to the contrast and texture modifications employed during training and/or testing time.

In most hospitals, the segmentations are done only for the baseline scans and not for the follow-up scans. This lack of GT segmentations creates a limitation for training the DL algorithms with follow-up scans. Since the performance of the models trained with ETZ only data is lower for the follow-up MRI compared to the performance for the planning MRI, we added the public data set to the training data and then evaluated the models trained with both ETZ and public data. The performance of the models on follow-up MRI images improved (when compared with the models trained with ETZ data only) by the addition of the public data to the training set and is comparable with other studies on BM segmentation on pre-treatment images [19, 21], suggesting this to be a viable solution to improve the model efficacy for the follow-ups in scenarios where the GT segmentations are lacking for follow-up MRI. This improvement in performance could be attributed to the increased heterogeneity introduced by the additional data in the training set, as well as the similarity in contrast enhancement (both single dose) between the public data in the training data set and the follow-up test data.

An interesting observation from this evaluation of nnU-Net and MedNeXt algorithms is that there is no remarkable difference in performance between the two algorithms. This is also evident from the performance comparison done by Roy et al. [35] which shows that MedNeXt comprehensively outperforms other algorithms for organ segmentation but in a more limited fashion for tumor segmentation. Considering that the training of MedNeXt algorithm takes additional time when compared to nnU-Net and the lack of a clear difference in performance between the two algorithms for BM segmentation, nnU-Net seems to be the favorable option. On the other hand, MedNext can be used when a marginal increase in performance is desired at the expense of increased training time.

To the best of our knowledge, the performance of the nnU-Net and MedNeXt algorithms exceeded the performance reported by other similar studies for segmentation of planning MRI. With the proposed approach to improve the performance of the models for the follow-up images, the algorithms hold promise as a valuable tool for clinicians for automated segmentation of MR scans, in diagnosis, treatment planning and treatment response evaluations during follow-ups. This study also demostrated a solution for the development for DL models in situations where the training data is sparse or not available. Also when there is large training data set, the addition of public data set could increase the heterogeneity of the training data and hence improve the model performance.

## Data Availability

The data used for this study is available at ETZ and is accessible after aproval from the ETZ Science office.

## Acknowledgments

We would like to acknowledge the support provided by Eline Verhaak for this research and thank her for helping us with the manually delineated ground truth for follow-up scans from the CAR study.

## Consent for publication

Not applicable.

## Contributions

All authors contributed to this research.

## Ethics approval

This study is part of the AI in Medical Imaging for novel Cancer User Support (AMICUS) project at Tilburg University. This project is approved by the Ethics Review Board at the Tilburg University.

## Consent to participate

Not applicable.

## List of abbreviations

SRS: Stereotactic Radiosurgery
BM: Brain metastases
MRI: Magnetic Resonance Imaging
CAR-Study: A Cognition And Radiation Study A
FU: Follow-up
DSC: Dice Similarity Coefficient
FNR: False Negative Rate
WBRT: Whole Brain Radiotherapy
DL: Deep learning
CNNs: Convolutional Neural Networks
CT: Computed Tomography scans
ETZ: Elisabeth-TweeSteden Hospital
GKRS: Gamma Knife Radiosurgery

